# The impact of the COVID-19 pandemic on the reporting of violence against children

**DOI:** 10.1101/2022.10.30.22281600

**Authors:** Camila dos Santos Souza Andrade, Maria da Conceição Nascimento Costa, Leny Alves Bonfim Trad, Marcio Santos da Natividade, Eliene dos Santos de Jesus, Rita de Cássia Oliveira de Carvalho-Sauer

**Affiliations:** Institute of Collective Health/Federal University of Bahia (ISC/UFBA); Retired professor of the Institute of Collective Health (ISC/UFBA); Researcher at the Center for Data Integration and Knowledge in Health (CIDACS), Gonçalo Moniz Institute, Oswaldo Cruz Foundation, Salvador, Bahia, Brazil; Professors of the Institute of Collective Health (ISC/UFBA); Coordinator at the Department of Health Information of the Municipal Secretariat of Health, Salvador, Bahia, Brazil; Technician at the East Regional Center for Health of the State Secretariat of Health of Bahia, Santo Antônio de Jesus, Bahia, Brazil

**Keywords:** violence against children, COVID-19, pandemic, impact, temporal trend study

## Abstract

**Objective:** To verify the impact (effect) of the COVID-19 pandemic on the rates of reporting of interpersonal violence against children aged 0-11 years old in Salvador, Bahia, from 2020 to 2021.

**Methods:** The study used two epidemiological approaches: a) temporal aggregation and b) an individual cross section, based on cases of interpersonal violence against children reported in SINAN from 2009 to 2021. Annual rates of reporting of interpersonal violence against children (per 10,000) and percentages were calculated according to different strata of each variable of interest. The temporal trend was analyzed using the simple linear regression method (R^2^=0.6955) applied to the rates from 2014 to 2019, the period in which they showed the most consistency.

**Results:** The rates of reporting of violence against children showed a large variation, with a mean of 4.7/10,000. In 2021, the rate was 7/10,000 (a 45.8% increase on the previous year). Regression analysis indicated a mean reduction of 0.337/10,000 a year, and expected rates of 4.62 and 4.28/10,000, respectively, for 2020 and 2021.

**Conclusion:** The occurrence of COVID-19 and, particularly, the increase in the number of reported cases of interpersonal violence against children in the second year of the pandemic in Bahia suggest that these events may be directly or indirectly related. More robust studies are needed to confirm this relationship.

## INTRODUCTION

Violence against children is a complex and multifaceted issue worldwide and represents an important public health problem with damaging effects on the child and society. Events of violence against children in the form of neglect, physical punishments, and psychological and sexual violence, among others, represent a phenomenon that is hard to understand and cause damaging psychosocial, cognitive, and behavioral repercussions in the individual, family, and collective environments^1^. There is thus the need for a deeper integration of efforts from the perspective of various disciplines, sectors, organizations, and communities. However, despite the comprehensiveness and advancement of the studies, the official statistics show a picture that requires greater investments from society focused on prevention and the protection of children.

A systematic review of the global prevalence of violence against children and adolescents aged from 2 to 14 and 15 to 17 years old, respectively, based on articles published after 2000, revealed that in Africa, Asia, and North America the minimum estimates for both age groups were close to or exceeded 50%, exceeding 30% in Latin America, while in Europe they tended to be lower^2^. In Brazil, a study based on cases of violence against children aged 0-9 years old reported by the public health services in 2011 indicated that 68.8% were aged up to 5 years old^3^.

Various factors are involved in the occurrence of this violence, such as unfavorable social conditions, unharmonious family relationships, a low educational level of the parents, and families in which the abusive or neglectful parents were abused or neglected in their childhood, among others^4^. Also, stress, insecurity, a breakdown of social and protection networks, and reduced access to services due to social isolation have been presented as alternative explanations for this phenomenon^5^. Regarding this last factor, the United Nations Children’s Fund (Unicef) indicates that more than 1.8 billion children experienced interruptions of violence prevention and response services due to COVID-19^6^. On the other hand, research conducted with data from Child Helpline International (CHI) revealed a 31% increase in the number of contacts for counseling received in the first six months of 2020 compared with the first six months of 2019, among the 40 child support lines, as well as a 35% increase in the first two quarters of 2020^7^.

In Bahia, the first case of COVID-19 was confirmed in March of 2020, some days after the confirmation of the first case in Brazil. As demonstrated by some authors, interruptions of violence prevention and response services occurred in different cities and states of the country, which may have affected the reporting of the event during the social isolation period^8^. However, there is not yet any information available on this issue in the state of Bahia. This study aimed to verify the impact of the COVID-19 pandemic on the evolution of the time series of rates of reporting of interpersonal violence against children aged 0-11 years old in Salvador, Bahia, in 2020 and 2021.

## METHODS

This retrospective study was conducted based on reported cases of interpersonal violence against children from 2009 to 2021, employing two epidemiological approaches: temporal aggregation, whose unit of analysis was the calendar year, and an individual cross section. The data relating to cases of interpersonal violence against children reported at the health units of the Unified Health System (SUS) were obtained from the Violence and Accidents Surveillance System (VIVA), in Salvador, the capital of the state of Bahia, made available by the Sub-coordinator of Health Information (SUIS) of that municipality. The study population comprised children residing in the capital aged 0 to 11 years old, the age group defined according to the age classification established by the Child and Adolescent Statute (ECA)^9^, provided by the Board of Epidemiological Surveillance of the Secretariat of Health of the State of Bahia (DIVEP/SESAB).

In the individual analysis, the following were employed: *variables relating to the victims* (sex, age group, schooling); *variables relating to the aggressor* (number involved, sex, alcohol use, and relationship with the victim); *characteristics of the violence* (typology, means of aggression, place of occurrence, recurrence); and *evolution and referrals to the health sector and other sectors*. The following were calculated: annual absolute frequencies, percentages, and rates (per 10,000) of reports of interpersonal violence against children in the periods from 2009 to 2021; and absolute frequencies and percentages according to the different strata of each variable aggregated for the total of the 2009-2019 and 2020-2021 periods, aiming to obtain greater robustness in the analysis, considering that the small annual number of cases reported in each stratum would result in instability of the indicators. For each variable, the strata without information were excluded. Next, a descriptive analysis of these was conducted. Differences between the proportions of the variables in the pre- and post-pandemic periods were tested by applying Pearson’s chi-squared test and trend test, considering a 5% significance level.

The impact of the COVID-19 pandemic on the trend of the rates of reporting of interpersonal violence against children was assessed by applying simple linear regression (point estimates and 95% confidence intervals). For a better adjustment of the model, we chose to consider the period from 2014, when the time series of the rate of reports showed a consistently decreasing trend (R^2^=0.6955). Next, the entire time series values, including the observed values and the values expected (predicted) in the pandemic period, were plotted on a graph, to enable a better comparison between the latter two, for 2020 and 2021.

The project of this study was approved by the Human Research Ethics Committee of the Collective Health Institute (ISC) of the Federal University of Bahia (UFBA) (CAAE 70601417.0.0000.5030).

## RESULTS

According to Table 1, from January of 2009 to December of 2021, the VIVA Surveillance System of Salvador reported the occurrence of 2753 cases of interpersonal violence against children aged 0 to 11 years old, a value that represents, on average, 6.6% of the number of cases reported for all ages. Higher proportions of reporting occurred in 2014 and 2015 (around 11%), in 2012 (9.4%), in 2018 (9.5%), and in 2021 (9.9%). In the study period, the mean rate of reporting was 4.7/10,000 children. It rose from 2.0/10,000 in 2009 to an average of 6.7/10,000 in 2014-2015. After that, an average of 5.4 cases were reported for every 10,000 children in 2016-2019, decreasing to 4.8/10,000 in 2020, and then rising to 7.0/10,000 in the following year. Therefore, a sharp inflection of the decreasing trend occurred, characterized by a 45.8% increase. Considering only the initial and final years of the rates of reports of interpersonal violence against children in the time series analyzed, a 250% increase is observed (from 2.0 in 2009 to 7.0/10,000 in 2021).

Among the cases with recorded information on each variable, in the 2009-2019 period, 57.2% of the victims were female, 56.6% were aged 0-5 years old, 90.9% were black and brown, respectively, 81.5% had a single perpetrator of the aggression, and 64.2% of the aggressors were male. It is also verified that 62.5% of the children’s aggressors were their own parents, followed by acquaintances of the victim (19.7%). Physical (42.0%) and sexual violence (31.0%) were the most frequent types of violence. In 2020-2021, 58.5% of the victims were female, 58.0% were aged 0-5 years old, 91.3% were black or brown girls, 80.7% had a single perpetrator of the aggression, 64.0% of the aggressors were male, and 81.2% did not involve alcohol use. It is also verified that 56.4% of the children’s aggressors were their own parents. The distribution of the race/skin color of the child victims of aggression and the aggressors’ sex did not show any significant statistical difference between the two periods analyzed (Table 2). Physical (33.0%) and sexual violence (28.2%) and neglect (27.4%) predominated.

In Table 3, it is verified that in 2014 and 2019 physical (46.1%) and sexual violence (28.6%) were the most frequent. Bodily force was the most used means for the aggression (52.2%), followed by objects (20.6%), while poisoning presented a 3.0% frequency. At home (74.1%) and in the street (17.8%) were the most frequent locations for the occurrence of the aggression. The health sector represented the main place the victims were referred to (58.1%), followed by police stations and the judiciary (24.6%). In 2020-2021, physical (33.0%) and sexual violence (28.2%) were also most frequent, by means of bodily force (39.2%) and objects (21.5%), occurring at home (88.5%) and in the street (9.5%), and primarily being referred to the healthcare network (42.0%) and police stations and the judiciary (21.4%). Poisoning occurred in 13.5% of the cases reported. Non-recurrence of the violence was most frequent both in 2009-2019 (60.9%) and in 2020-2021 (58.0%), with this being the only variable analyzed that did not show a significant difference between the respective frequencies recorded for both periods.

A simple linear regression analysis applied for 2014 to 2019, the period in which the trend of the rate of reports of interpersonal violence against children was most consistent (p=0.039; R^2^=0.6955), indicated a mean reduction of 0.337/10,000 every year. The values expected for the pandemic period were 4.62 (95%CI: 3.41; 5.83) for 2020, thus being within the expected values, and 4.28/10,000 (95%CI: 2.79; 5.77) for 2021 (Figure 2021); that is, in the latter year the aforementioned rate (7.0/10,000) exceeded the expected value, representing an excess of 64%.

## DISCUSSION

The results of the present study reveal that in Salvador, Bahia, in 2020, the first year of the COVID-19 pandemic, the value observed for the rate of reporting of interpersonal violence against children aged up to 11 years old and resident in the municipality was within the expected limits, indicating that this indicator may not have been impacted by the pandemic. However, in the following year, this health event produced a strong impact on that rate, given that the observed value of 7.0/10,000 represented a 45.8% increase in relation to the previous year (4.8/10,000) and a 65% excess in relation the expected value of 4.3 (3.8-5.8/10,000). During the whole study period (2009-2021), the rate of reporting of interpersonal violence against the children analyzed presented a large variation (mean value of 4.7/10,000) and, for that reason, the predicted values were obtained based on the data from 2014 to 2019, when a consistently decreasing mean trend was observed, in the order of 0.337/10,000 a year. In both the pre- and post-pandemic periods, the most frequent victims were female, black, and aged 5 years old at most. The parents were the main aggressors, being mostly male, and the aggression generally occurred in the child’s home. Physical violence followed by sexual violence and neglect predominated, with the latter presenting 73.4% growth in 2020-2021 (it rose from 15.8% in 2009-2019 to 27.4% in 2020-2021). The 350% increase observed in violence through poisoning draws attention, which rose from 3.0% in 2009-2019 to 13.5% in 2020-2021. Only the child’s race/skin color, the aggressor’s sex, and the recurrence of the aggression of the child did not present significant differences between the frequencies recorded in both periods analyzed.

It is known that children’s vulnerability is sometimes exacerbated by a lack of understanding of the totality of the social chaos caused by the pandemic and sometimes because they are the object of numerous cases of violence both by their family members and/or the absence of the state. In more vulnerable contexts, material and social deficiencies and unsatisfied everyday needs mean that affective relationships are often imbued with conflicts^10^. In his explanatory proposal for intra-family violence, the systemic theory, Bronfenbrenner (1979)^11^ reinforces the possible explanations for the increase in the violence in the confinement period by elucidating that this type of violence communicates with the social fabric; that is, that socioeconomic, environmental, and cultural factors, among others, act as stressors and in turn increase the possibility of violent events. These facts, allied with the intensification of the activities in the surveillance sector in compliance with the current health protocols for the municipality of Salvador during the pandemic, may explain the increase in reports of violence against children observed in our study in 2021.

On the other hand, the reduction in the aforementioned reports observed in our study in 2020, the first year of the COVID-19 pandemic in Brazil, corroborates with results of other investigations carried out in the south region of the country^12,8^ and in US states^13^ and translates as intense underreporting, primarily due to the reduction in the activities of health, education, and social welfare professionals, who offer the children and their families support through identification and referrals to the protective services in cases of interpersonal violence against children^6,14,15^. Therefore, it is important to highlight that the reduction in reporting does not mean a reduction in this type of violence. On the contrary, the confinement caused much tension in family relationships, due to the context of much uncertainty and insecurity^8^.

The increase in the reports of violence observed in our study from 2009 to 2013 was also found for children and adolescents in Rio Grande do Sul in this same period^16^ and according to the authors of that investigation it resulted from the implementation of the technical guidance of Ordinance n. 104/2011^17^, which made violence an event that should be obligatorily reported by health professionals^18^. However, the literature indicates certain obstacles for professionals to file these reports, such as weaknesses in the institutional routine regarding legal protection, fear of judicial involvement, fear of the aggressor, and disbelief in the accountability by the competent bodies^19^. These factors may have contributed to the decline in reports in the years from 2016 to 2019.

The panorama of violence outside the pandemic context experienced by the children included in this study supports the idea that violence against children is a practice rooted in the family environment, caused by a family aggressor or acquaintance^20,21,22^. It has been related with cultural questions, gender issues (machismo, patriarchy, authoritarianism), and power as a form of conflict resolution^21^. The intersection of racial and gender questions also means that black girls suffer from double inequality and discrimination at the same time and in the same space^23^, a phenomenon they are subjected to and that may accompany them in other age groups.

As Magalhães et al. (2017)^24^ mention, the practice of violence against children is justified as an permitted educational resource, considered to be appropriate, common, culturally accepted, and therefore socially reproduced. As opposed to the protective role, in various social conditions the family is shown to be a privileged space for the expression of violence. However, it should be stressed that despite its direct relationship with social inequality, the violence against children practiced in the family environment does not involve a particular social class, but primarily the sociocultural construction of the interpersonal relationships of the actors involved^25^.

The limitations of this study include the incompleteness of the fields of the VIVA form, the underreporting of cases, and little or almost no capturing of cases in private services. Combined with that, the reports may only refer to serious cases of violence that required medical assistance. All of these factors have an impact on the magnitude of the phenomenon, generating underestimated indicators. The underreporting of violence results in a fragmented and disconnected perception of it by the various sectors of society. In turn, the low quality of the information means manifestations of violence in the infant segment are practically invisible to managers, hindering the planning of actions and public policies^26^. Its incompleteness compromises the execution of more robust analyses, often impeding the identification of risk and protective factors^27^.

The exacerbation of interpersonal violence against children in the second year of the pandemic unfortunately reinforced/revealed the timid engagement by the state in comprehensive child protection actions and projects, as well as the difficulties in relying on the support of the school, guardianship councils, and other social centers. This context further exposed the vulnerability of the children, reinforcing the need to strengthen health services to improve/increase the number of reports and the quality of the information provided on the form, as well as of connecting the families and their children in a situation of violence with the network for prevention, care, and the guarantee of rights.

## Data Availability

All data produced in the present work are contained in the manuscript

## Authorship

CSSA and MCNC developed the conception and design of the study. CSSA, MCNC, LABT, MSN, ESJ, and RCOCS contributed to the analysis and interpretation of the data. CSSA, MCNC, and RCOCS wrote the first version of the article. All the authors participated in the critical review of the manuscript and approved the version to be published.

## Funding

None

**Figure 1.**
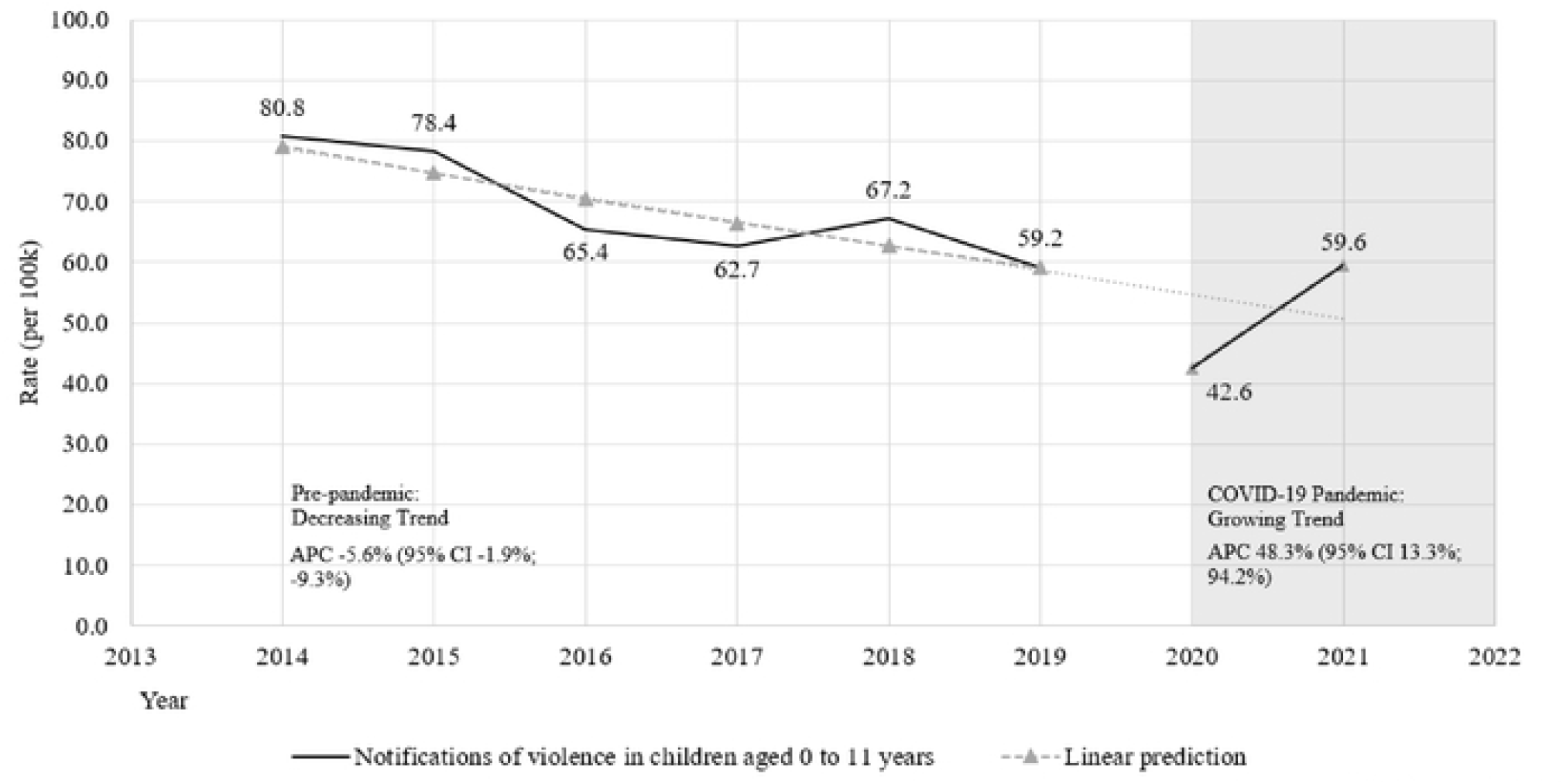
Interrupted time series, trend and annual percentage change in reporting rates of interpersonal violence in children aged 0-11 years in the pre-pandemic (2014-2019) and pandemic (2020-2021) periods of COVID-19. Salvador, Bahia, Brazil, 2014-2021. APC = Annual pcrccntagedumge.3

